# Seroprevalence, waning, and correlates of anti-SARS-CoV-2 IgG antibodies in Tyrol, Austria: Large-scale study of 35,193 blood donors conducted between June 2020 and September 2021

**DOI:** 10.1101/2021.12.27.21268456

**Authors:** Anita Siller, Lisa Seekircher, Gregor A. Wachter, Manfred Astl, Lena Tschiderer, Bernhard Pfeifer, Manfred Gaber, Harald Schennach, Peter Willeit

## Abstract

**Background:** There is uncertainty about the seroprevalence of anti-SARS-CoV-2 antibodies in the general population of Austria, and about the extent to which antibodies elicited by vaccination or infection wane over time.

**Aim:** To estimate seroprevalence, waning, and correlates of anti-SARS-CoV-2 IgG antibodies in the Federal State of Tyrol, Austria.

**Methods:** We conducted a seroepidemiological study between June 2020 and September 2021, enrolling blood donors aged 18-70 years across Tyrol, Austria (participation rate 84.0%). We analysed serum samples for antibodies against spike or nucleocapsid proteins of SARS-CoV-2 with Abbott SARS-CoV-2 IgG assays.

**Results:** We performed 47,363 serological tests among 35,193 individuals (median age 43.1 years [IQR: 29.3-53.7], 45.3% women, 10.0% with prior SARS-CoV-2 infection). Seroprevalence increased from 3.4% (95% CI: 2.8-4.2%) in June 2020 to 82.7% (95% CI: 81.4-83.8%) in September 2021, largely due to vaccination. Anti-spike IgG seroprevalence was 99.6% (99.4-99.7%) among fully vaccinated individuals, 90.4% (88.8-91.7%) among unvaccinated with prior infection, and 11.5% (10.8-12.3%) among unvaccinated without known prior infection. Anti-spike IgG levels were reduced by 44.0% (34.9-51.7%) at 5-6 months compared to 0-3 months after infection. In fully vaccinated individuals, they decreased by 31.7% (29.4-33.9%) per month. In multivariable adjusted analyses, both seropositivity among unvaccinated and antibody levels among fully vaccinated individuals were higher at young age (<25 years), higher with a known prior infection, and lower in current smokers.

**Conclusion:** Seroprevalence in Tyrol increased to 82.7% in September 2021, with the bulk of seropositivity stemming from vaccination. Antibody levels substantially and gradually declined after vaccination or infection.

## Introduction

The severe acute respiratory syndrome coronavirus type 2 (SARS-CoV-2) pandemic has been the major public health concern worldwide since December 2019. By the end of November 2021, more than 260 million SARS-CoV-2 infections and 5.2 million related deaths have been reported globally [1]. At the same time, approximately 44% of the world population has been fully vaccinated and 55% has been partially vaccinated against SARS-CoV-2 [1]. As a considerable proportion of populations has already been infected or vaccinated, seroepidemiological studies are important to quantify reliably the seroprevalence of anti-SARS-CoV-2 antibodies at the population level and thereby inform future decision making of governments and health authorities.

There are several studies in different European countries that have assessed the trajectories of anti-SARS-CoV-2 seroprevalence in the general population over time, including studies in blood donors and household samples in Germany [2] and the UK [3,4]. In Austria, however, previous seroprevalence studies have been restricted to the time before vaccinations against SARS-CoV-2 were licensed [5–7] and, hence, data on seroprevalence in the vaccination-era is lacking. Moreover, there are scarce data on the rate at which anti-SARS-CoV-2 antibody levels wane after an infection and after vaccination [8,9].

To address these uncertainties, we conducted a large-scale seroepidemiological study among blood donors in the Federal State of Tyrol, Austria. Our aims were threefold. First, to reliably determine anti-SARS-CoV-2 IgG seroprevalence for each month between June 2020 and September 2021 and according to the nine districts of Tyrol. Second, to characterise antibody dynamics after SARS-CoV-2 infection and after vaccination. Third, to quantify reliably any differences in anti-SARS-CoV-2 IgG antibodies across relevant population subgroups.

## Methods

Results are reported in accordance with the Strengthening the Reporting of Observational studies in Epidemiology (STROBE) guidelines (**Supplementary Table S1**).

### Study design and participants

We herein report on a retrospective cohort study conducted in blood donors in the Federal State of Tyrol in Austria. Study participants were recruited between 8 June 2020 and 30 September 2021 at 145 different blood donation events spread across all districts of Tyrol. Individuals were eligible for inclusion if they (i) were aged between 18 and 70 years; (ii) were permanent residents in Tyrol; (iii) and fulfilled the general requirements for donating blood, including being in a healthy state (e.g. free of malignant diseases, auto-immune diseases, or infectious diseases). Participants were asked to complete a questionnaire that included information on lifestyle factors (i.e. height, weight, and smoking) and prior SARS-CoV-2 infection (i.e. date of diagnosis and method of detection). In addition, data on age, sex, and SARS-CoV-2 vaccination were collected routinely as part of every blood donation. For the latter, individuals were classified as “fully vaccinated” if they received (i) two doses of the BNT162b2, mRNA-1273, or ChAdOx1-S vaccine; (ii) one dose of the Ad26.COV2.S vaccine; or (iii) one dose of any vaccine in case of a past PCR-confirmed SARS-CoV-2 infection. Of 41,941 eligible individuals, a total of 35,214 individuals took part in the study (participation rate 84.0%). After excluding individuals without a laboratory result (n=17) and without information on their vaccination status (n=4), 35,193 individuals contributed to our analysis.

### Ethical statement

The present study was approved by the ethics committee of the Medical University of Innsbruck (no. 1352/2021). The planning, conduct, and reporting of the study is in line with the Declaration of Helsinki, as revised in 2013.

### Laboratory analyses

Serum samples were drawn, cooled at 4°C, and normally processed within ≤30 hours centrally at the laboratory of the Central Institute for Blood Transfusion and Immunology of the University Hospital in Innsbruck, Austria. Two types of chemiluminescent microparticle immunoassays were used in our study. First, between 8 June 2020 and 31 March 2021, we assessed samples for anti-SARS-CoV-2 IgG antibodies targeting the nucleocapsid protein (“anti-N IgG”) using the Abbott SARS-CoV-2 IgG chemiluminescent microparticle immunoassay analysed on the Alinity i instrument (Abbott Ireland, Sligo, Ireland). According to the manufacturer, the assay has a sensitivity of 100% (95% confidence interval [CI]: 95.89-100%) at ≥14 days after COVID-19 onset (post-symptom onset), and a specificity of 99.63% (99.05-99.90%) [10]. Second, between 19 March 2021 and 30 September 2021, we assessed samples for anti-SARS-CoV-2 IgG antibodies targeting the spike protein (“anti-S IgG”) using the Abbott SARS-CoV-2 IgG II chemiluminescent microparticle immunoassay analysed on the Alinity i instrument. The manufacturer’s recommended cut-off value for positivity of ≥7.1 Binding Antibody Units per millilitre (BAU/mL) has a sensitivity of 99.35% (96.44-99.97%) at ≥15 days after COVID-19 onset (post-symptom onset) and a specificity of 99.60% (99.22-99.80%) [11]. The switch to the anti-S IgG assay was done because the assay allows quantification of absolute antibody levels (in BAU/mL) and detects anti-SARS-CoV-2 IgG antibodies elicited via vaccination.

### Statistical analyses

The primary analysis quantified seroprevalence of anti-SARS-CoV-2 IgG antibodies for each of the months between June 2020 and September 2021 and is presented together with Agresti-Coull 95% CIs. Serological tests in August and September 2020 were excluded because they were considered as non-representative samples (e.g. 71.4% in Ischgl [12]). Because the offer to test for anti-SARS-CoV-2 IgG antibodies as part of a blood donation may have mobilised individuals and may thereby have introduced some selection bias, we conducted a sensitivity analysis restricted to individuals with repeat donations since October 2017. Furthermore, we conducted an age- and sex-standardisation using the structure of the total population in Tyrol aged 18-70 years to enhance generalisability of the study estimates [13]. Finally, seroprevalence was also estimated separately for the nine districts of Tyrol and – to facilitate interpretation – is presented together with (i) cumulative SARS-CoV-2 incidence calculated from public data [14] and (ii) state-wide vaccine coverage provided to the authors by the Corona Operations Centre of the Crises and Disaster Management Team of the Government of Tyrol.

We conducted several subsidiary analyses that focused on the time period with anti-S IgG measurements. First, we quantified seroprevalence in population subgroups defined by vaccination status and prior SARS-CoV-2 infection and tested for differences in antibody levels using linear regression. Second, to investigate differences by population subgroups, we fitted multivariable regression models that included the variables age (<25 vs. ≥25 years), sex (males vs. females), smoking (current vs. never/ex-smokers), body mass index (≥25 vs. <25 kg/m^2^), prior SARS-CoV-2 infection (yes vs. no) based on complete case analysis. In specific, we used (i) a generalised estimating equation with a logit link function, a binomial distribution family, and an independent variance structure to test for differences in seroprevalence among unvaccinated individuals, and (ii) a linear mixed model with a random intercept to test for differences in antibody levels after full vaccination. Third, we investigated the stability of anti-S IgG antibody levels after SARS-CoV-2 infection and after vaccination using linear mixed models with random intercepts that incorporated all available repeat measurements. Furthermore, to assess the shape of association with time since SARS-CoV-2 infection, we entered restricted cubic splines with three equidistant knots spread across the range of time since infection into the linear mixed model. Because the distribution of anti-S IgG antibody levels was skewed, we used log-transformed values in all regression models and back-transformed results for presentation. P values ≤0.05 were deemed as statistically significant and all statistical test were two-sided. Analyses were carried out with Stata 15.1 and R 4.1.0.

## Results

### Study population

**Table 1** summarises the characteristics of the 35,193 participants enrolled in our study. Median age was 43.1 years (interquartile range [IQR] 29.3-53.7), 45.3% were female, and 17.8% were current smokers. Participants had a mean body mass index of 25.2 kg/m^2^ (standard deviation [SD] 3.9) and 10.0% reported having had a SARS-CoV-2 infection in the past. Of the 35,193 participants, 24,483 (69.6%) had a measurement of anti-N IgG and 19,792 (56.2%) had a measurement of anti-S IgG. Repeat measurements of SARS-CoV-2 antibodies were available in 9,735 participants (27.7%) over a median follow-up duration of 6.3 months (IQR 5.4-9.4). In total, 47,363 serological tests were performed.

**Table 1.** Characteristics of participants enrolled in our study, Tyrol, Austria, June 2020-September 2021 (n=35,193).

| | Total no. | No. (%), mean ± SD, median (IQR), or range |
| --- | --- | --- |
| **Baseline data** |  |  |
| Date of baseline – range | 35,193 | 8 Jun 2020-30 Sep 2021 |
| Age in years – median (IQR) | 35,193 | 43.1 (29.3-53.7) |
| Female sex – no. (%) | 35,193 | 15,950 (45.3%) |
| Current smoker – no. (%) | 28,265 | 5,035 (17.8%) |
| Body mass index in kg/m<sup>2</sup> – mean ± SD | 28,176 | 25.2 ± 3.9 |
| Prior SARS-CoV-2 infection – no. (%) | 31,039 | 3,113 (10.0%) |
| First donation since Oct 2017 – no. (%) | 35,193 | 13,832 (39.3%) |
| **Availability of SARS-CoV-2 antibody data** |  |  |
| Participants with anti-N IgG – no. (%) | 35,193 | 24,483 (69.6%) |
| Participants with anti-S IgG – no. (%) | 35,193 | 19,792 (56.2%) |
| **Repeat donations during study** |  |  |
| Participants with $\geq 2$ donations – no. (%) | 35,193 | 9,735 (27.7%) |
| Follow-up duration in months – median (IQR) | 9,735 | 6.3 (5.4-9.4) |
| Abbreviations: IQR, interquartile range; SD, standard deviation. |  |  |

### Seroprevalence of anti-SARS-CoV-2 IgG antibodies by time and by region

**Figure 1** shows the evolution of anti-SARS-CoV-2 IgG seroprevalence during the course of our study. Seroprevalence of anti-N IgG antibodies was 3.4% (95% CI: 2.8-4.2%) in June 2020, peaked in January 2021 at 17.1% (16.0-18.3%), and declined slightly to 14.0% (13.0-15.1%) in March 2021. Seroprevalence of anti-S IgG antibodies increased from 29.9% (27.5-32.4%) in March 2021 (i.e. when the anti-S assay was introduced in our study) to 82.7% (81.4-83.8%) in September 2021. As depicted in **Figure 1**, this increase was largely due to the vaccination rollout, with a seroprevalence among individuals that remained unvaccinated of 17.1% (15.2-19.3%) in March 2021 and 7.4% (6.5-8.2%) in September 2021. Among unvaccinated participants with concomitant information on both anti-SARS-CoV-2 IgG antibodies (n=1,582), test results agreed for 90.7% (i.e. 201 both positive, 1,234 both negative), were positive for anti-S IgG only in 8.7% (n=138), and were positive for anti-N IgG only in 0.6% (n=9). In sensitivity analyses restricted to participants who had already donated blood leading up to the study (**Supplementary Table S2**), seroprevalences were slightly higher than in the principal analysis in the final five months of the study (e.g. 84.4% [82.8-85.8%] in September 2021). The seroprevalence estimates standardised to the age- and sex-structure of the population in Tyrol are provided in **Supplementary Table S2** (e.g. 82.6% [81.3-83.8%] in September 2021).

**Figure 1.**
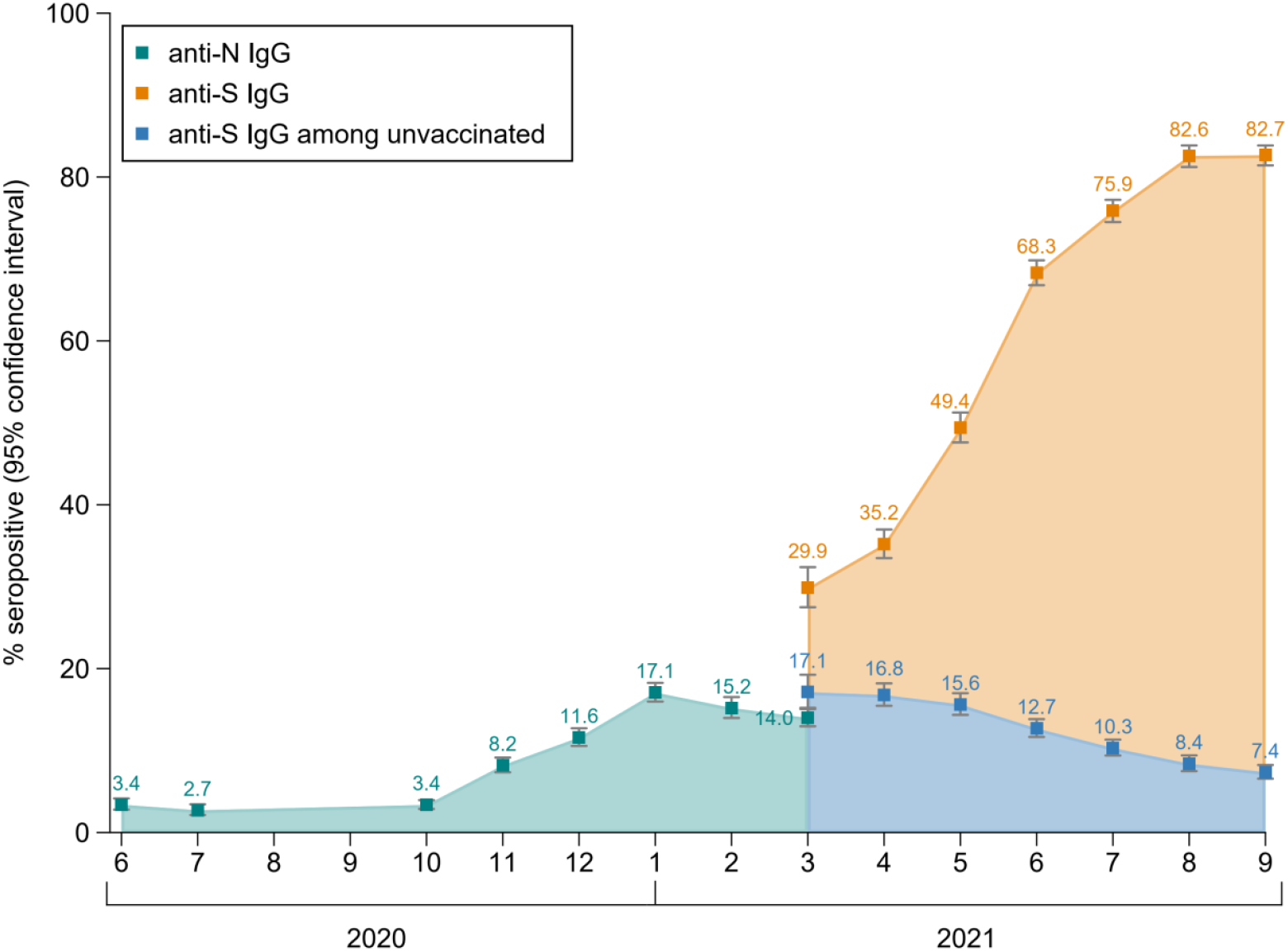
Seroprevalence of anti-SARS-CoV-2 IgG antibodies in Tyrolean blood donors aged 18-70 years, Tyrol, Austria, June 2020-September 2021 (n=35,193). The analysis involved data on 47,363 measurements taken from 35,193 individuals.

**Figure 2** compares regional differences in cumulative SARS-CoV-2 incidence and vaccination coverage in the total population of Tyrol with regional differences in the proportion of our study population that seroconverted up to the period from July to September 2021. We observed the lowest seroprevalence in the district of Lienz (74.3% [70.8-77.4%]), which was also the district with the lowest vaccine coverage in Tyrol with 54.4% being fully vaccinated by end-September 2021. In contrast, we observed the highest seroprevalence in the district of Schwaz (87.6% [85.8-89.3%]). In this district, vaccine coverage increased sharply in April 2021 from 6% to 56% due to a population-wide rapid rollout vaccination campaign and remained the highest across Tyrol since then. Furthermore, the district of Schwaz had the highest cumulative incidence of documented SARS-CoV-2 infections in Tyrol up to April 2021 (10.6%).

**Figure 2.**
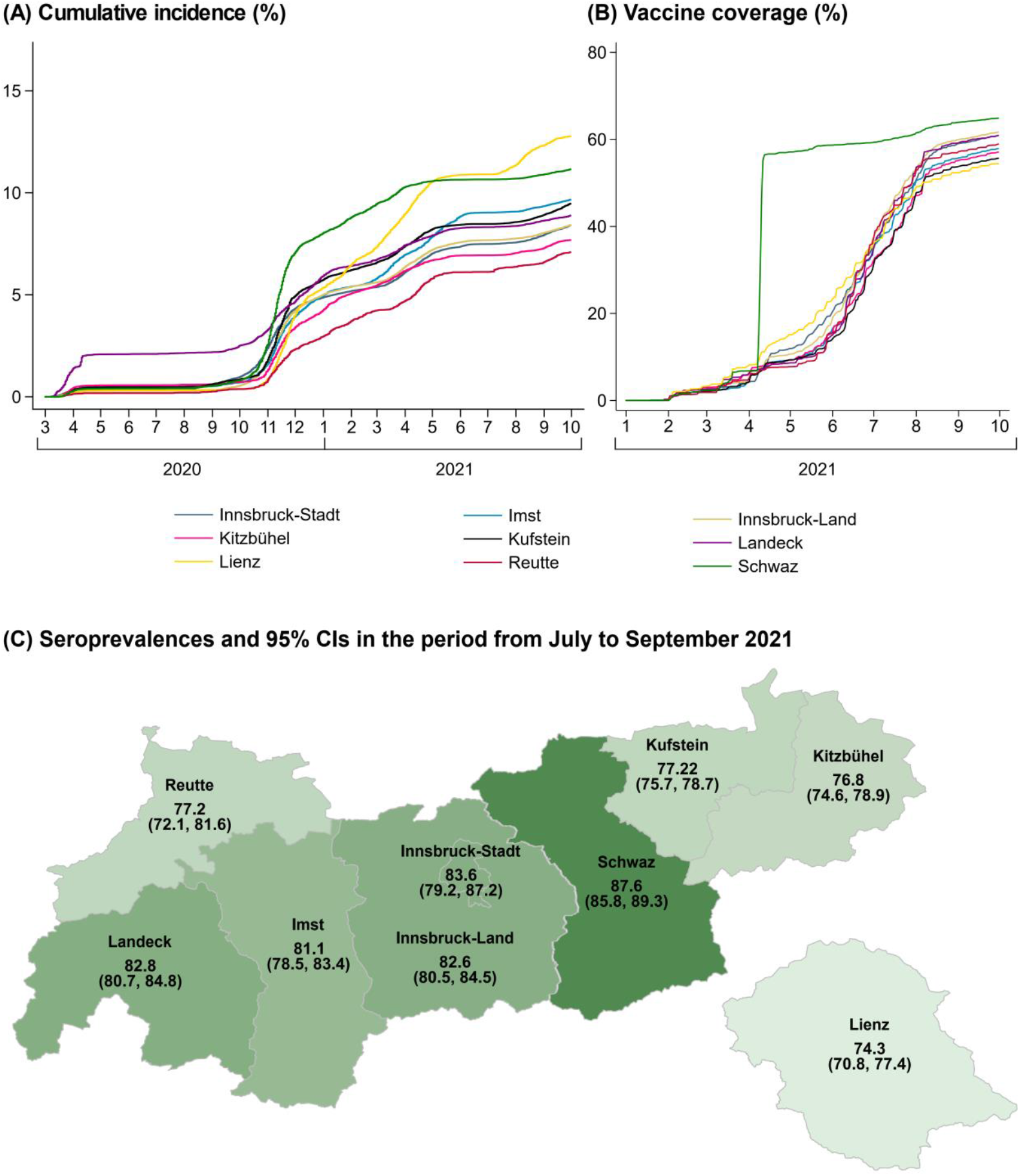
Regional differences in cumulative SARS-CoV-2 incidence and vaccine coverage in the total population of Tyrol (Panel A and B) and seroprevalence of anti-S IgG antibodies in the blood donors aged 18-70 years enrolled in our study (Panel C), Tyrol, Austria, March 2020-September 2021 (n=760,105) and July-September 2021 (n=10,632). Abbreviation: CI, confidence interval.

### Seroprevalence of anti-S IgG antibodies by vaccination status and prior SARS-CoV-2 infection

To assess seroprevalence by vaccination status and prior SARS-CoV-2 infection, we analysed data on 19,792 individuals with a measurement of anti-S IgG (**Table 2**). Among participants vaccinated against SARS-CoV-2, seroprevalence was 79.9% (95% CI: 78.3-81.4%) in those classified as partially vaccinated and 99.6% (99.4-99.7%) in fully vaccinated participants. Among unvaccinated participants, seroprevalence was 90.4% (88.8-91.7%) in those reporting a prior SARS-CoV-2 infection (a median of 5.6 months prior to the assessment [IQR 3.9–7.4]) and 11.5% (10.8-12.3%) in those reporting not to have had a prior SARS-CoV-2 infection. The antibody level differed significantly between partially vaccinated participants (median 70 BAU/mL [IQR 15-259]), fully vaccinated participants (757 [265-1,829]), unvaccinated participants with prior SARS-CoV-2 infection (38 [17-87]), and unvaccinated participants without prior SARS-CoV-2 infection (0.1 [0.0-0.5]) (all P values for difference<0.001).

**Table 2.** Seroprevalence of anti-S IgG antibodies in vaccinated and unvaccinated study participants, Tyrol, Austria, March-September 2021 (total n=19,792).

|  | **Seropositive**<br>**/ total no.** | **% seropositive (95%**<br>**CI)** | **Median level (IQR) in**<br>**BAU/mL** |
| --- | --- | --- | --- |
| **Vaccinated** |  |  |  |
| Partially vaccinated | 2,048 / 2,563 | 79.9 (78.3-81.4) | 70 (15-259) |
| Fully vaccinated<sup>a</sup> | 8,098 / 8,133 | 99.6 (99.4-99.7) | 757 (265-1,829) |
| **Unvaccinated** |  |  |  |
| Prior SARS-CoV-2 infection | 1,445 / 1,599 | 90.4 (88.8-91.7) | 38 (17-87) |
| No prior SARS-CoV-2 infection | 815 / 7,070 | 11.5 (10.8-12.3) | 0.1 (0.0-0.5) |
| Missing information | 111 / 427 | 26.0 (22.1-30.4) | 0.2 (0.0-9.1) |

Abbreviations: BAU, binding antibody units; CI, confidence interval; IQR, interquartile range.
<sup>a</sup>Individuals were classified as fully vaccinated if they had received two doses of the BNT162b2, mRNA-1273, or ChAdOx1-S vaccine or one dose of the Ad26.COV2.S vaccine or they had recovered from COVID-19 and had received one dose of any vaccine. The analysis focused on each donor's earliest survey with anti-S IgG information.

### Cross-sectional correlates of anti-S IgG seroprevalence and antibody levels

**Table 3** shows the results of the multivariable regression models we fitted to investigate cross-sectional correlates of anti-S IgG antibodies. First, in an analysis of the group of unvaccinated participants, the odds ratio for being seropositive for anti-S IgG antibodies was 2.06 for participants aged <25 years (95% CI: 1.52-2.78), 0.39 for current smokers (0.27-0.56), 1.31 for participants with a body mass index of 25 kg/m^2^ or higher (1.02-1.69), and 64.81 for participants with prior SARS-CoV-2 infection (48.33-86.92). Compared to this analysis of the period from July to September 2021, supplementary analyses of earlier periods of the study yielded largely similar findings (**Supplementary Figure S1**). Second, in an analysis of the group of fully vaccinated participants, anti-S IgG antibody levels were 51.9% higher in participants aged <25 years (37.8-67.4%) and 129.3% higher in those with prior SARS-CoV-2 infection (109.0-151.6%), 8.0% lower in males (2.1-13.6%), and 10.6% lower in current smokers (2.7-17.9%).

**Table 3.** Cross-sectional correlates of seroprevalence and level of anti-S IgG antibodies, Tyrol, Austria, July-September 2021 (n=2,684) and March-September 2021 (n=7,701).

|  | Seroprevalence among unvaccinated<sup>a</sup><br>(n=2,684) |  | Anti-S IgG level among fully vaccinated<sup>b</sup><br>(n=7,701) |  |
| --- | --- | --- | --- | --- |
|  | % seropositive<br>(95% CI) | Multivariable adjusted<sup>c</sup><br>odds ratio (95% CI)<br>vs. reference | Level in BAU/mL,<br>median (IQR) | Multivariable adjusted<sup>c</sup><br>% difference (95% CI)<br>vs. reference |
| **Age groups** |  |  |  |  |
| 25 years or older | 29.4 (27.5-31.3) | [Reference] | 675 (249-1,617) | [Reference] |
| <25 years | 39.6 (35.2-44.3) | 2.06 (1.52-2.78) | 1,233 (350-2,781) | +51.9 (37.8 to 67.4) |
| **Sex** |  |  |  |  |
| Female | 30.2 (27.7-32.9) | [Reference] | 776 (264-1,803) | [Reference] |
| Male | 31.7 (29.4-34.1) | 1.20 (0.94-1.54) | 676 (250-1,680) | -8.0 (-13.6 to -2.1) |
| **Smoking status** |  |  |  |  |
| Never/ex-smoker | 33.8 (31.8-35.8) | [Reference] | 753 (263-1,794) | [Reference] |
| Current smoker | 19.5 (16.3-23.1) | 0.39 (0.27-0.56) | 580 (220-1,486) | -10.6 (-17.9 to -2.7) |
| **Body mass index** |  |  |  |  |
| <25 kg/m<sup>2</sup> | 30.4 (28.2-32.8) | [Reference] | 743 (254-1,772) | [Reference] |
| 25 kg/m<sup>2</sup> or higher | 31.8 (29.2-34.6) | 1.31 (1.02-1.69) | 697 (258-1,712) | +4.6 (-1.8 to 11.4) |
| **Prior SARS-CoV-2 infection** |  |  |  |  |
| No | 12.7 (11.3-14.2) | [Reference] | 633 (229-1,559) | [Reference] |
| Yes | 89.6 (87.0-91.8) | 64.81 (48.33-86.92) | 1,491 (668-2,986) | +129.3 (109.0 to 151.6) |

Abbreviations: IQR, interquartile range; SD standard deviation. <sup>a</sup>The analysis of seroprevalence among unvaccinated individuals focuses on the period from July to September 2021, thereby reflecting infections that occurred up to this period. Results for earlier periods are provided in **Supplementary Figure S1**.
<sup>b</sup>Individuals were classified as fully vaccinated if they had received two doses of the BNT162b2, mRNA-1273, or ChAdOx1-S vaccine or one dose of the Ad26.COV2.S vaccine or they had recovered from COVID-19 and had received one dose of any vaccine. <sup>c</sup>All variables shown in this table were included in the multivariable adjusted models.

### Waning of anti-S IgG antibody levels after a SARS-CoV-2 infection and after vaccination

We next quantified the decrease of anti-S IgG antibody levels over time after a person had a SARS-CoV-2 infection or was vaccinated. In an analysis of 1,455 unvaccinated participants with 1,573 repeat measurements (**Figure 3**), we observed substantial waning of anti-S IgG antibody levels most notably in the first six months after infection. For instance, compared to the median anti-S IgG levels of 60 BAU/mL (IQR 30-130) at month 0-3 after an infection, anti-S IgG levels were 44.0% lower at month 5-6 (95% CI: 34.9-51.7%) and 58.8% lower at month 10 or later (50.7-65.6%) (**Figure 3B**). Finally, in an analysis of 269 fully vaccinated participants with 2 or more visits resulting in a total of 548 repeat measurements, we observed mean attrition rate in anti-S IgG antibody levels of 31.7% per month (95% CI: 29.4-33.9%).

**Figure 3.**
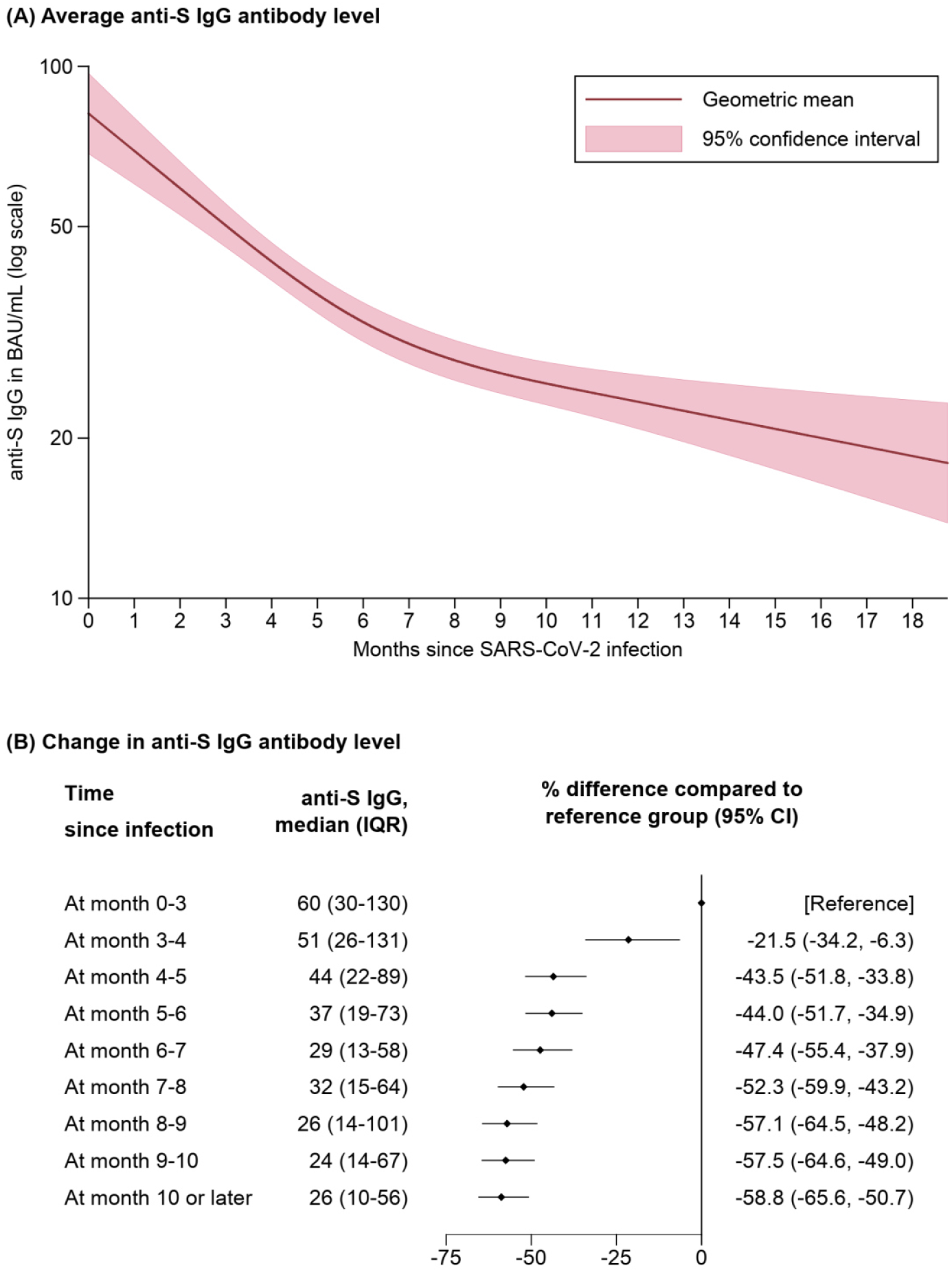
Waning of anti-S IgG antibody levels after a SARS-CoV-2 infection among unvaccinated blood donors, Tyrol, Austria, March-September 2021 (n= 1,455a). Abbreviations: CI, confidence interval; IQR, interquartile range. ^a^1,455 participants are included in the analysis and 113 participants provided repeat anti-S IgG antibody levels. The analysis presented in Panel (A) used a mixed model based on restricted cubic splines with three equidistant knots around the range of infection duration, with anti-S IgG antibody values being log-transformed for analysis and estimated coefficients being exponentiated to reflect geometric mean levels at different time points.

## Discussion

The present large-scale study reports on the seroprevalence of anti-SARS-CoV-2 IgG antibodies between summer 2020 and autumn 2021 in 35,193 healthy individuals aged 18-70 years recruited at blood donation events throughout all districts of Tyrol, Austria. In September 2021, seroprevalence was 82.7%, was largely attributable to vaccination rather than past infection, and varied across districts (e.g. 87.6% in the district of Schwaz vs. 74.3% in the district of Lienz). Furthermore, on top of the anticipated differences in seroprevalence and antibody levels by vaccination status and prior SARS-CoV-2 infection, we identified cross-sectional correlations with young age, known prior infection, and smoking that withstood multivariable adjustment. Finally, by incorporating the repeat measurements taken in our longitudinal study, we reliably quantified the progressive waning of antibody levels, occurring after a SARS-CoV-2 infection and after vaccination.

Our study provides much anticipated seroprevalence data that will inform upcoming decisions in the control of the pandemic in Austria. All prior population-based seroepidemiological studies in Austria have been conducted in 2020 and hence before vaccination against SARS-CoV-2 was rolled out to the public. These include a first report from our cohort [5] that revealed a seroprevalence of 3.1% (95% CI: 2.7-3.6%) in June to September 2020 in Tyrol and another study [6] in blood donors in four other Federal States of Austria that showed a seroprevalence of 2.5% (2.2-2.7%) in June to December 2020 in four other Federal States of Austria. Furthermore, in November 2020, a nationwide study involving individuals selected based on household sampling found a seroprevalence of 3.1% (95% CI: 2.2-4.0%) [7]. In contrast to the scarce data in Austria, studies in other European countries have documented the built-up of seroprevalence in the year 2021, e.g. an increase from 3% to 17% by April among German blood donors [2], an increase from 14% to 61% by May among participants of the English REACT-2 study [3], and an increase from 8-11% to 91-93% by September across the countries of the United Kingdom [4]. In our study, by September 2021, we found an increase of seroprevalence to 82.7% (95% CI: 81.4-83.8%), which was mainly due to the high vaccination uptake in this age group (18-70 years), while less than a tenth was attributable to seropositivity induced by infection only. Our study also revealed regional differences across the nine districts of the Tyrol, which were broadly in line with known vaccine coverages and the number of incident SARS-CoV-2 cases documented by the health authorities in the total population. Finally, the age- and sex-distribution in our cohort of blood donors and the general population was well aligned and thereby standardisation of seroprevalence for these demographic variables yielded very similar results.

The remarkably high immunogenicity in fully vaccinated individuals is another important finding, with 99.6% exhibiting anti-S IgG antibodies at the time of blood donation. Partially vaccinated individuals were seropositive only in 79.9% and, when compared to the fully vaccinated group, also had – on average – a more than 10-fold lower absolute level of anti-S IgG. Our data on immunogenicity which are largely based on vaccination with BNT162b2 (i.e. 72% of doses in Tyrol by the end of the study [14], compared to 16% with ChAdOx1-S and 10% with mRNA-1273) are in close agreement with the time trends following receipt of first and second dose vaccination previously reported for mRNA COVID-19 vaccines [15–17]. Given the limited phenotypic information available in our study, we could not investigate in detail why a small fraction of participants (0.4%) were non-responders to full vaccination, but different immunocompromising conditions are associated with a lack in immunogenicity [18]. Nevertheless, substantially higher anti-S IgG levels were visible in younger individuals and individuals with prior SARS-CoV-2 infection, while lower anti-S IgG levels were detected among male participants and smokers. Furthermore, by capitalising on the longitudinal data available in our study, we were able to precisely quantify the rate of anti-S IgG antibody waning after vaccination. The overall reduction of 31.7% per month (95% CI: 29.4-33.9%) we observed in our study closely agrees with a previous longitudinal study that investigated waning of antibody levels in 1,647 health care workers after the second dose of a mRNA COVID-19 vaccine [8], but is slightly less than the 18.3-fold decrease reported in 3,808 health care workers six months after receipt of dose two of the BNT162b2 vaccine [9], The minimal antibody level however that is required for preventing a SARS-CoV-2 infection or for preventing severe COVID-19 as well as the importance of cell-mediated immunity is still unclear [19,20]. Nevertheless, our findings support the notion that effectiveness of COVID-19 vaccines is reduced after 4-6 months [21–23] and that a booster dose is needed to reinstate the protection conferred by the vaccines [19].

We also conducted a range of analyses focusing on seroprevalence in unvaccinated individuals with or without a prior SARS-CoV-2 infection. Participants with prior SARS-CoV-2 infection were seropositive in 90.4% and – consistent with prior data [16,17] – had a lower antibody level than fully vaccinated participants. A novel contribution of our study is that we were able to characterise the shape of antibody waning after a SARS-CoV-2 infection. In specific, our analyses identified a steep decline of antibody levels in the first six months post-infection (resulting in a 44.0% reduction in total at month 5-6) and a slower decline in the subsequent months. Furthermore, our study revealed that 11.5% of people reporting not to have had SARS-CoV-2 infection actually had been infected unknowingly. The finding in our multivariable adjusted analyses that younger and overweight/obese individuals were more commonly seropositive, while smokers were less commonly seropositive, could stem from (i) differing degrees of exposure and subsequent infection rates or (ii) differing immune responses and antibody dynamics following an infection [5,24].

A much discussed concept related to seroprevalence studies is so-called ‘herd immunity’ or ‘population immunity’, i.e. a state in which a sufficient proportion of the population is immune to a pathogen to prevent the spread of the pathogen in the entire population including among those that lack immunity. Based on the basic reproduction number of 2.5-3.5 of the virus wild type [25], it was initially estimated that 60-72% immunity is required to reach herd immunity for SARS-CoV-2.

However, this threshold has been constantly shifted upwards as more infectious SARS-CoV-2 variants [26] or potential immune escape variants have emerged [27], and as vaccine-induced immunity is not distributed evenly across populations (i.e. differences between countries, difference by age groups, vaccine hesitancy), thereby maintaining high susceptibility to infection in specific groups of individuals [28]. Furthermore, while vaccines against SARS-CoV-2 are highly effective in preventing symptomatic COVID-19, they do not offer complete protection from infection and may not block forward-transmission entirely [29,30], thereby undermining a pillar of herd immunity. While there is uncertainty about future developments of the pandemic, in an expert consultation paper, we have previously outlined key determinants and possible courses of the pandemic over the next years [28].

The study we presented herein has several important strengths and limitations. With data on 35,193 participants with 47,363 analysed samples, our study was adequately powered to reliably quantify time- and region-specific seroprevalence. Furthermore, because a subset of participants donated blood repeatedly, we gained important insight into the dynamics of antibody levels over time. Also, while the eligibility criteria for blood donation restricted our study sample to healthy individuals aged 18-70 years, the age- and sex-structure of our study sample was in close agreement with the general population, thereby supporting the generalisability of our findings to these age groups. Still, when interpreting our seroprevalence estimates, it is crucial to take into account that vaccine coverage among children and adolescents who constitute 17.4% of the population in Tyrol is much lower and therefore seroprevalence across all age groups is expected to be lower. Limitations of our study include (i) the availability of anti-N IgG antibody measurements only up to March 2021 (precluding a clear differentiation of infection- and vaccination-induced seropositivity) and (ii) lack of detailed data on the vaccination regimen (i.e. vaccination dates and vaccine type) because of time constraints at the blood donation centres.

## Conclusion

In conclusion, seroprevalence of anti-SARS-CoV-2 antibodies increased from 3.4% in June 2020 to 82.7% in September 2021, with the bulk of seropositivity stemming from vaccination. This study also highlights a substantial gradual decline of antibody levels after vaccination or after SARS-CoV-2 infection. Furthermore, it illustrates that blood donors are well suited as an easily accessible study group for seroepidemiological studies and that they can help to identify local outbreaks of infectious diseases.

## Data Availability

Data on COVID-19 cases in districts of Tyrol and on the age and sex structure of the population in Tyrol are publicly available. Tabular data on the blood donor cohort can be requested from the corresponding authors by researchers who submit a methodologically sound proposal (including a statistical analysis plan); participant-level data on the blood donor cohort cannot be shared due to regulatory restrictions.

https://www.data.gv.at/katalog/dataset/4b71eb3d-7d55-4967-b80d-91a3f220b60c

http://www.statistik.at/web_de/statistiken/menschen_und_gesellschaft/bevoelkerung/bevoelkerungsstruktur/bevoelkerung_nach_alter_geschlecht/index.html

## Conflict of interest statement

The authors report no conflicts of interest in relation to this study.

## Funding statement

The study was supported by the Federal State of Tyrol and the Tirol Kliniken GmbH. The funders had no role in study design, data collection, data analysis, or writing of the report. The corresponding authors had full access to all data in the study and had final responsibility for the decision to submit for publication.

## Acknowledgements

The authors thank all employees of the infection serology laboratory of the Central Institute for Blood Transfusion and Immunology, Tirol Kliniken GmbH, namely Barbara Schennach, Elfriede Lanser, Julia Penz, Andrea Schiestl, Sabrina Schmid, and Michaela Szabo. We also thank Katharina Lerchster, Barbara Schlögl, and Helga Egger for processing of questionnaire data and all employees of the blood donation service of the Tyrolean Red Cross.

## Authors’ contributions

AS, GAW, BP, HS, and PW designed the study. AS and HS coordinated the study. MG and HS supervised the enrolment of the blood donors. AS and GAW supervised the processing of study questionnaires. MA, LS, and LT performed data management. LS and LT performed data cleaning and statistical analysis. LS, AS, and PW drafted the manuscript. All authors critically revised the manuscript and agreed to be accountable for all aspects of the work.

## Notes

### Competing Interest Statement

The authors have declared no competing interest.

### Author Declarations

The ethics committee of the Medical University of Innsbruck gave ethical approval for this work (no. 1352/2021).

## References

1. Ritchie H, Mathieu E, Rodés-Guirao L, Appel C, Giattino C, Ortiz-Ospina E, et al. Coronavirus Pandemic (COVID-19). Available from: https://ourworldindata.org/coronavirus. Accessed at: 13 Dec 2021.

2. Robert Koch Institut. Serologische Untersuchungen von Blutspenden auf Antikörper gegen SARS-CoV-2 (SeBluCo-Studie). Available from: https://www.rki.de/DE/Content/InfAZ/N/Neuartiges_Coronavirus/Projekte_RKI/SeBluCo_Zwischenbericht.html. Accessed at: 10 Dec 2021.

3. Ward H, Whitaker M, Tang SN, Atchison C, Darzi A, Donnelly CA, et al. Vaccine uptake and SARS-CoV-2 antibody prevalence among 207,337 adults during May 2021 in England: REACT-2 study. medRxiv. 2021. https://doi.org/10.1101/2021.07.14.21260497.preprint.

4. Office for National Statistics. Antibodies against coronavirus (COVID-19). Available from: https://www.ons.gov.uk/peoplepopulationandcommunity/healthandsocialcare/conditionsanddiseases/articles/coronaviruscovid19latestinsights/antibodies. Accessed at: 26 Nov 2021.

5. Siller A, Wachter GA, Neururer S, Pfeifer B, Astl M, Borena W, et al. Prevalence of SARS-CoV-2 antibodies in healthy blood donors from the state of Tyrol, Austria, in summer 2020. Wien Klin Wochenschr. 2021. https://doi.org/10.1007/s00508-021-01963-3.

6. Weidner L, Nunhofer V, Jungbauer C, Hoeggerl AD, Grüner L, Grabmer C, et al. Seroprevalence of anti-SARS-CoV-2 total antibody is higher in younger Austrian blood donors. Infection. 2021. https://doi.org/10.1007/s15010-021-01639-0.

7. Statistik Austria. Mehr als die Hälfte der SARS-CoV-2-Infektionen kurz vor dem zweiten Lockdown sind behördlich nicht erfasst. Available from: https://www.statistik.at/web_de/presse/124846.html. Accessed at: 10 Dec 2021.

8. Steensels D, Pierlet N, Penders J, Mesotten D, Heylen L. Comparison of SARS-CoV-2 Antibody Response Following Vaccination With BNT162b2 and mRNA-1273. JAMA. 2021;326:1533–5. https://doi.org/10.1001/jama.2021.15125.

9. Levin EG, Lustig Y, Cohen C, Fluss R, Indenbaum V, Amit S, et al. Waning Immune Humoral Response to BNT162b2 Covid-19 Vaccine over 6 Months. N Engl J Med. 2021. https://doi.org/10.1056/NEJMoa2114583.

10. Abbott Alinity i SARS-CoV-2 IgG Instructions for Use. December 2020.

11. Abbott Alinity i SARS-CoV-2 IgG II Quant Instructions for Use. April 2021.

12. Borena W, Bánki Z, Bates K, Winner H, Riepler L, Rössler A, et al. Persistence of immunity to SARS-CoV-2 over time in the ski resort Ischgl. EBioMedicine. 2021;70:103534. https://doi.org/10.1016/j.ebiom.2021.103534.

13. Statistik Austria. Bevölkerung nach Alter und Geschlecht. Available from: http://www.statistik.at/web_de/statistiken/menschen_und_gesellschaft/bevoelkerung/bevoelkerungsstruktur/bevoelkerung_nach_alter_geschlecht/index.html. Accessed at: 09 Dec 2021.

14. data.gv.at - Open Data Österreich. Katalog COVID-19: Zeitliche Darstellung von Daten zu Covid19-Fällen je Bezirk. Available from: https://www.data.gv.at/covid-19/. Accessed at: 07 Dec 2021.

15. Walsh EE, Frenck RW, Falsey AR, Kitchin N, Absalon J, Gurtman A, et al. Safety and Immunogenicity of Two RNA-Based Covid-19 Vaccine Candidates. N Engl J Med. 2020;383:2439–50. https://doi.org/10.1056/NEJMoa2027906.

16. Naranbhai V, Garcia-Beltran WF, Chang CC, Mairena CB, Thierauf JC, Kirkpatrick G, et al. Comparative immunogenicity and effectiveness of mRNA-1273, BNT162b2 and Ad26.COV2.S COVID-19 vaccines. medRxiv. 2021. https://doi.org/10.1101/2021.07.18.21260732. preprint.

17. Anderson EJ, Rouphael NG, Widge AT, Jackson LA, Roberts PC, Makhene M, et al. Safety and Immunogenicity of SARS-CoV-2 mRNA-1273 Vaccine in Older Adults. N Engl J Med. 2020;383:2427–38. https://doi.org/10.1056/NEJMoa2028436.

18. Rahav G, Lustig Y, Lavee J, Ohad B, Magen H, Hod T, et al. BNT162b2 mRNA COVID-19 vaccination in immunocompromised patients: A prospective cohort study. EClinicalMedicine. 2021;41:101158. https://doi.org/10.1016/j.eclinm.2021.101158.

19. Burckhardt RM, Dennehy JJ, Poon LLM, Saif LJ, Enquist LW. Are COVID-19 Vaccine Boosters Needed? The Science behind Boosters. J Virol. 2021:JVI0197321. https://doi.org/10.1128/JVI.01973-21.

20. Turner JS, O’Halloran JA, Kalaidina E, Kim W, Schmitz AJ, Zhou JQ, et al. SARS-CoV-2 mRNA vaccines induce persistent human germinal centre responses. Nature. 2021;596:109–13. https://doi.org/10.1038/s41586-021-03738-2.

21. Tartof SY, Slezak JM, Fischer H, Hong V, Ackerson BK, Ranasinghe ON, et al. Effectiveness of mRNA BNT162b2 COVID-19 vaccine up to 6 months in a large integrated health system in the USA: a retrospective cohort study. Lancet. 2021;398:1407–16. https://doi.org/10.1016/S0140-6736(21)02183-8.

22. Thomas SJ, Moreira ED, Kitchin N, Absalon J, Gurtman A, Lockhart S, et al. Safety and Efficacy of the BNT162b2 mRNA Covid-19 Vaccine through 6 Months. N Engl J Med. 2021;385:1761–73. https://doi.org/10.1056/NEJMoa2110345.

23. Rosenberg ES, Dorabawila V, Easton D, Bauer UE, Kumar J, Hoen R, et al. Covid-19 Vaccine Effectiveness in New York State. N Engl J Med. 2021. https://doi.org/10.1056/NEJMoa2116063.

24. Nomura Y, Sawahata M, Nakamura Y, Kurihara M, Koike R, Katsube O, et al. Age and Smoking Predict Antibody Titres at 3 Months after the Second Dose of the BNT162b2 COVID-19 Vaccine. Vaccines (Basel). 2021;9. https://doi.org/10.3390/vaccines9091042.

25. Billah MA, Miah MM, Khan MN. Reproductive number of coronavirus: A systematic review and meta-analysis based on global level evidence. PLoS One. 2020;15:e0242128. https://doi.org/10.1371/journal.pone.0242128.

26. Campbell F, Archer B, Laurenson-Schafer H, Jinnai Y, Konings F, Batra N, et al. Increased transmissibility and global spread of SARS-CoV-2 variants of concern as at June 2021. Euro Surveill. 2021;26. https://doi.org/10.2807/1560-7917.ES.2021.26.24.2100509.

27. Pulliam JRC, van Schalkwyk C, Govender N, Gottberg A von, Cohen C, Groome MJ, et al. Increased risk of SARS-CoV-2 reinfection associated with emergence of the Omicron variant in South Africa. [place unknown]: [publisher unknown]; 2021.

28. Iftekhar EN, Priesemann V, Balling R, Bauer S, Beutels P, Calero Valdez A, et al. A look into the future of the COVID-19 pandemic in Europe: an expert consultation. Lancet Reg Health Eur. 2021;8:100185. https://doi.org/10.1016/j.lanepe.2021.100185.

29. Singanayagam A, Hakki S, Dunning J, Madon KJ, Crone MA, Koycheva A, et al. Community transmission and viral load kinetics of the SARS-CoV-2 delta (B.1.617.2) variant in vaccinated and unvaccinated individuals in the UK: a prospective, longitudinal, cohort study. Lancet Infect Dis. 2021. https://doi.org/10.1016/S1473-3099(21)00648-4.

30. Shah ASV, Gribben C, Bishop J, Hanlon P, Caldwell D, Wood R, et al. Effect of Vaccination on Transmission of SARS-CoV-2. N Engl J Med. 2021;385:1718–20. https://doi.org/10.1056/NEJMc2106757.

