## Supplementary Material for "Seroprevalence, waning, and correlates of anti-SARS-CoV-2 IgG antibodies in Tyrol, Austria: Large-scale study of 35,193 blood donors conducted between June 2020 and September 2021"

### Supplementary tables

#### Table S1. STROBE checklist.

| | Item No | Recommendation | Page No |
|---|---|---|---|
| Title and abstract | 1 | (*a*) Indicate the study's design with a commonly used term in the title or the abstract | 1 |
| | | (*b*) Provide in the abstract an informative and balanced summary of what was done and what was found | 2 |
| **Introduction** | | | |
| Background/ rationale | 2 | Explain the scientific background and rationale for the investigation being reported | 3 |
| Objectives | 3 | State specific objectives, including any prespecified hypotheses | 3 |
| **Methods** | | | |
| Study design | 4 | Present key elements of study design early in the paper | 3-4 |
| Setting | 5 | Describe the setting, locations, and relevant dates, including periods of recruitment, exposure, follow-up, and data collection | 3-4 |
| Participants | 6 | (*a*) Give the eligibility criteria, and the sources and methods of selection of participants. Describe methods of follow-up | 3-4 |
| | | (*b*) For matched studies, give matching criteria and number of exposed and unexposed | NA |
| Variables | 7 | Clearly define all outcomes, exposures, predictors, potential confounders, and effect modifiers. Give diagnostic criteria, if applicable | 3-5 |
| Data sources/ measurement | 8* | For each variable of interest, give sources of data and details of methods of assessment (measurement). Describe comparability of assessment methods if there is more than one group | 3-5 |
| Bias | 9 | Describe any efforts to address potential sources of bias | 4-5 |
| Study size | 10 | Explain how the study size was arrived at | 3-4 |
| Quantitative variables | 11 | Explain how quantitative variables were handled in the analyses. If applicable, describe which groupings were chosen and why | 4-5 |
| Statistical methods | 12 | (*a*) Describe all statistical methods, including those used to control for confounding | 4-5 |
| | | (*b*) Describe any methods used to examine subgroups and interactions | 4-5 |
| | | (*c*) Explain how missing data were addressed | 4-5 |
| | | (*d*) If applicable, explain how loss to follow-up was addressed | NA |
| | | (*e*) Describe any sensitivity analyses | 4-5 |
| **Results** | | | |
| Participants | 13* | (a) Report numbers of individuals at each stage of study—eg numbers potentially eligible, examined for eligibility, confirmed eligible, included in the study, completing follow-up, and analyzed | 6-10 |
| | | (b) Give reasons for non-participation at each stage | 6-10 |
| | | (c) Consider use of a flow diagram | NA |
| Descriptive data | 14* | (a) Give characteristics of study participants (eg demographic, clinical, social) and information on exposures and potential confounders | 6, Table 1 |
| | | (b) Indicate number of participants with missing data for each variable of interest | Table 1 |
| | | (c) Summarize follow-up time (eg, average and total amount) | Table 1 |
| Outcome data | 15* | Report numbers of outcome events or summary measures over time | 6-10 |
| Main results | 16 | (a) Give unadjusted estimates and, if applicable, confounder-adjusted estimates and their precision (eg, 95% confidence interval). Make clear which confounders were adjusted for and why they were included | 6-10 |
| | | (b) Report category boundaries when continuous variables were categorized | 6-10 |
| | | (c) If relevant, consider translating estimates of relative risk into absolute risk for a meaningful time period | NA |
| Other analyses | 17 | Report other analyses done—eg analyses of subgroups and interactions, and sensitivity analyses | 7-10 |
| **Discussion** | | | |
| Key results | 18 | Summarize key results with reference to study objectives | 10 |
| Limitations | 19 | Discuss limitations of the study, taking into account sources of potential bias or imprecision. Discuss both direction and magnitude of any potential bias | 12-13 |
| Interpretation | 20 | Give a cautious overall interpretation of results considering objectives, limitations, multiplicity of analyses, results from similar studies, and other relevant evidence | 10-13 |
| Generalizability | 21 | Discuss the generalizability (external validity) of the study results | 12-13 |
| **Other information** | | | |
| Funding | 22 | Give the source of funding and the role of the funders for the present study and, if applicable, for the original study on which the present article is based | 2;5 |

**Table S2. Seroprevalences in all participants (principal analysis), in participants with repeated donations since October 2017, and in all participants with age- and sex-standardisation across the total population of Tyrol.**

| Month | % seropositive (95% CI) | | |
|---|---|---|---|
| | **All participants (n=35,193)** | **Participants who had already donated blood leading up to the study[a] (n=21,361)** | **All participants (n=35,193) with age- and sex-standardisation across the total population of Tyrol[b]** |
| **Anti-N IgG** | | | |
| June 2020 | 3.4 (2.8-4.2) | 2.5 (2.0-3.3) | 3.3 (2.6-4.0) |
| July 2020 | 2.7 (2.1-3.4) | 2.6 (1.9-3.4) | 2.6 (2.0-3.3) |
| October 2020 | 3.4 (2.9-4.0) | 3.1 (2.5-3.7) | 3.3 (2.7-3.9) |
| November 2020 | 8.2 (7.3-9.1) | 7.4 (6.4-8.5) | 7.8 (6.9-8.7) |
| December 2020 | 11.6 (10.6- 12.7) | 11.4 (10.1-12.9) | 11.2 (10.1-12.3) |
| January 2021 | 17.1 (16.0-18.3) | 16.9 (15.5-18.4) | 16.8 (15.7-18.0) |
| February 2021 | 15.2 (14.0-16.5) | 16.5 (14.9-18.2) | 15.3 (14.0-16.7) |
| March 2021 | 14.0 (13.0-15.1) | 14.1 (12.9-15.5) | 14.3 (13.5-15.2) |
| **Anti-S IgG** | | | |
| March 2021 | 29.9 (27.5-32.4) | 29.4 (26.5-32.5) | 29.1 (26.6-31.6) |
| April 2021 | 35.2 (33.5-37.0) | 36.3 (34.2-38.5) | 35.6 (33.7-37.5) |
| Mai 2021 | 49.4 (47.6-51.3) | 55.1 (52.8-57.3) | 52.2 (50.4-54.1) |
| June 2021 | 68.3 (66.8-69.8) | 73.5 (71.6-75.3) | 68.5 (67.0-70.1) |
| July 2021 | 75.9 (74.5-77.2) | 79.9 (78.3-81.5) | 76.0 (74.6-77.4) |
| August 2021 | 82.6 (81.2-83.9) | 84.9 (83.2-86.4) | 82.7 (81.4-84.0) |
| September 2021 | 82.7 (81.4-83.8) | 84.4 (82.8-85.8) | 82.6 (81.3-83.8) |

[a]To define this subgroup of the study population, the period from October 2017 to study baseline was considered. [b]Direct age- and sex standardisation was applied by using age (categories 18-30, >30-40, >40-50, >50-60, >60-70 years) and sex structured data of the population of the Federal State of Tyrol in Austria as standard population as of 1 January 2021 from the Statistik Austria. Seroprevalence estimates by age groups and sex are presented in **Supplementary Figure S2**.

#### Supplementary figures

**Figure S1. Seroprevalence of anti-S IgG antibodies among unvaccinated and of anti-N IgG antibodies for different time periods and across different population subgroups, Tyrol, Austria, June 2020-September 2021 (n=35,193).**

| Month | % seropositive[a] | Adjusted[b] odds ratio (95% CI) | P-value |
|---|---|---|---|
| **Age <25 vs. ≥25** | | | |
| Jun-Jul 2020 | 4.6 / 2.8 | 1.70 (1.15, 2.50) | 0.008 |
| Oct-Dec 2020 | 8.9 / 7.2 | 1.33 (1.09, 1.63) | 0.005 |
| Jan-Mar 2021 | 16.7 / 16.5 | 1.05 (0.91, 1.21) | 0.474 |
| Apr-Jun 2021 | 25.4 / 24.8 | 1.06 (0.90, 1.25) | 0.469 |
| Jul-Sep 2021 | 39.6 / 29.4 | 1.67 (1.34, 2.08) | <0.001 |
| **Male vs. female** | | | |
| Jun-Jul 2020 | 3.3 / 2.9 | 1.19 (0.87, 1.63) | 0.287 |
| Oct-Dec 2020 | 7.4 / 7.4 | 0.95 (0.82, 1.10) | 0.502 |
| Jan-Mar 2021 | 16.6 / 16.4 | 0.97 (0.87, 1.08) | 0.607 |
| Apr-Jun 2021 | 25.4 / 24.4 | 1.04 (0.92, 1.19) | 0.529 |
| Jul-Sep 2021 | 31.8 / 30.0 | 1.06 (0.89, 1.26) | 0.494 |
| **Current vs. never/ex-smoker** | | | |
| Oct-Dec 2020 | 4.1 / 8.1 | 0.48 (0.37, 0.61) | <0.001 |
| Jan-Mar 2021 | 8.8 / 18.1 | 0.43 (0.36, 0.52) | <0.001 |
| Apr-Jun 2021 | 17.2 / 26.7 | 0.57 (0.47, 0.68) | <0.001 |
| Jul-Sep 2021 | 19.5 / 33.7 | 0.47 (0.37, 0.59) | <0.001 |
| **BMI ≥25 vs. <25** | | | |
| Oct-Dec 2020 | 8.0 / 6.9 | 1.24 (1.07, 1.44) | 0.005 |
| Jan-Mar 2021 | 17.5 / 15.7 | 1.16 (1.04, 1.30) | 0.008 |
| Apr-Jun 2021 | 25.5 / 24.5 | 1.07 (0.93, 1.22) | 0.346 |
| Jul-Sep 2021 | 31.8 / 30.4 | 1.14 (0.96, 1.36) | 0.133 |

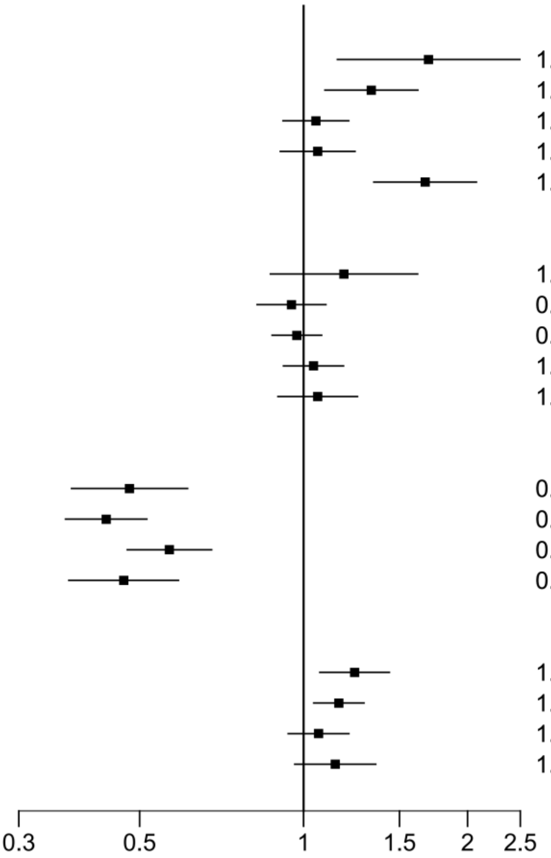


Abbreviations: BMI, body mass index; CI, confidence interval. [a]The reference group is depicted on the right side. [b]Adjusted for all the variables shown in this figure (age <25 vs. ≥25 years), male vs. female sex, current vs. never/ex-smoker, and body mass index (≥25 vs. <25 kg/m²).

**Figure S2. Seroprevalence of anti-SARS-CoV-2 IgG antibodies by sex and age group, Tyrol, Austria, June 2020-September 2021 (n=35,193).**

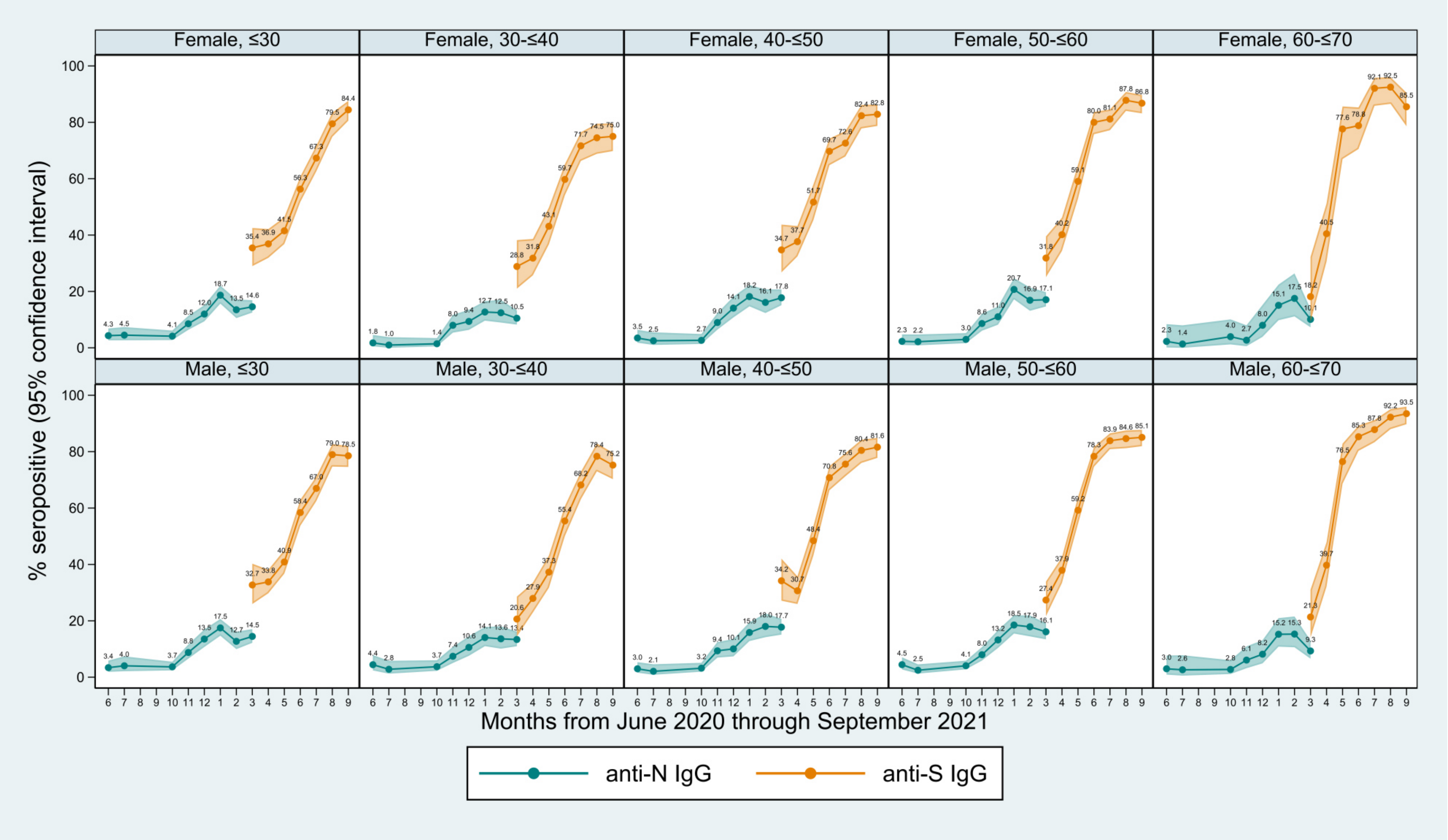
